# Sex Differences in Cognitive Performance in Alzheimer’s Disease: Insights from the ADAS–Cog-13

**DOI:** 10.1101/2025.07.08.25331133

**Authors:** Mohan Liu, Khushi Bhatt, Mia F. Giaever, Babak A. Ardekani, Chelsea Reichert Plaska, Elham Ghanbarian, the Alzheimer’s Disease Neuroimaging Initiative

**Affiliations:** School of Global Public Health, New York University, NY, USA; Department of Neurology, University of California, Irvine, CA, USA; Middlebury College, VT, USA; Center for Biomedical Imaging and Neuromodulation, The Nathan S. Kline Institute for Psychiatric Research, Orangeburg, NY, USA; Department of Psychiatry, NYU Grossman School of Medicine, NY, USA

**Keywords:** Alzheimer’s disease, sex, ADAS-Cog-13, hippocampus, brain, MRI

## Abstract

Sex differences in Alzheimer’s disease (AD) are well recognized, yet their impact on cognitive assessment remains underexplored. We examined sex differences in the 13-item AD Assessment Scale–Cognitive Subscale (ADAS-Cog-13) separately within cognitively normal (CN), and AD patients, controlling for hippocampal atrophy. CN women performed significantly better than men on the total score, word recall, naming, and word recognition. No sex differences were observed within the AD group on the total score, but males performed better in delayed word recall. These findings underscore the need to account for sex when interpreting cognitive test results, especially in preclinical stages of AD.

## 1. Introduction

The most common cause of dementia worldwide is Alzheimer’s Disease (AD), which affects more than 50 million people. Approximately two-thirds of those with AD clinical diagnoses are females.^1,2^ Studies have shown that there are sex differences in the progression and clinical presentation of AD, possibly as a result of hormones, genetics, and other biological differences.^3^ Regarding clinical progression, females with mild cognitive impairment (MCI), an intermediate stage in the AD continuum, progress to late-stage AD twice as fast as men.^4,5,6^ While females show better cognitive outcomes than men in the early stages of the disease,^7,8,9,10^ as the disease progresses, they show faster decline.^11,12^

Previous studies have shown sex differences in cognitive performance among both cognitively normal (CN) individuals and patients with AD. Females typically outperform males on verbal memory tasks across the lifespan.^13,14,15^ These differences are also observed in early AD and may contribute to delayed or less frequent diagnosis of AD in women in earlier stages of the disease.^16,17^ Sex differences in the clinical presentation of AD may compromise the reliability of standard neuropsychological assessments, which often overlook inherent sex-based variations in cognitive performance.^18^ Therefore, examining sex differences in cognitive test performance is essential for accurate interpretation and diagnosis.

The Alzheimer’s Disease Assessment Scale–Cognitive Subscale (ADAS-Cog-13) is a comprehensive 13-question metric that measures global cognition using features of language, orientation, praxis, and memory.^19^ Studies on sex differences in ADAS-Cog-13 performance have been inconclusive, making it important to further investigate the sex differences in cognitive assessments.^20,21^ Because evaluation of cognitive abilities contributes to more accurate diagnosis in the AD continuum, identifying the role of sex differences in these evaluations is crucial. Furthermore, no studies have yet compared sex-based differences in ADAS questionnaire sub-scores, which may reveal more domain-specific sex differences in cognitive tasks. In the present study, we explored whether there are sex differences in overall ADAS-Cog-13 scores and sub-scores in CN individuals and in AD patients while controlling for hippocampal volumetric integrity, age, and education.

## 2. Methods

### 2.1. Participants

All data used in the preparation of this manuscript were obtained from the Alzheimer’s Disease Neuroimaging Initiative (ADNI) database (adni.loni.usc.edu). The ADNI was launched in 2003 as a public-private partnership, led by Principal Investigator Michael W. Weiner, MD. The primary goal of ADNI has been to test whether serial magnetic resonance imaging (MRI), positron emission tomography, other biological markers, and clinical and neuropsychological assessment can be combined to measure the progression of MCI and early AD. Refer to www.adni-info.org for more details regarding the ADNI procedures and diagnostic criteria.

For the current study, we followed the ADNI2 protocol to determine the diagnostic and inclusion criteria for participants. Subjects classified within the ADNI2 dataset as CN (n=656) or mild AD (n=193) were included in this study.

### 2.2. Hippocampal parenchymal fraction (HPF)

For all participants, a 3D high-resolution T1-weighted structural MRI was used to measure hippocampal volumetric integrity. Hippocampal parenchymal fraction (HPF) is the ratio of preserved hippocampal tissue to the total volume in a standardized hippocampal volume of interest (VOI). A lower HPF indicates greater hippocampal degeneration. HPF for the left and right hemispheres were calculated using the KAIBA algorithm (v.3.0) of the Automatic Registration Toolbox (ART).^22,23^ The bilateral HPF values were averaged to obtain a single measure of hippocampal volumetric integrity.

### 2.3. The 13-item Alzheimer’s Disease Assessment Scale–Cognitive Subscale (ADAS-Cog-13)

The ADAS-Cog-13 is a 13-item cognitive assessment that evaluates performance across multiple domains, including memory, language, visuospatial ability, and attention.^24, 25^ Total scores range from 0 to 85, with higher scores indicating greater cognitive impairment. In this study, we used ADAS-Cog-13 data from the ADNI database, and analyses were based on both the total summary score and each of the 13 individual items separately. This approach allowed us to assess not only overall cognitive performance, but also domain-specific impairments associated with diagnostic status and other covariates of interest.

### 2.4. Statistical analysis

Independent samples t-tests were used to compare basic population characteristics between males and females. The following multiple regression model was subsequently employed to analyze the associations between ADAS scores in CN and AD groups:

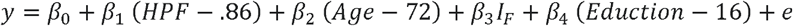

In this model, the intercept (*β*_0_) indicates the baseline ADAS-Cog-13 score of a 72-year-old, male subject with an HPF of 86% and 16 years of education. These demographic factors were chosen, because they were the marginal means of all study participants. The model coefficients *β*_1_, *β*_2_, *β*_3_, and *β*_4_ represent effects of the predictor variables HPF, age, female sex, and education, respectively. These represent the expected change in the intercept ADAS-Cog-13 score per unit increase in the respective predictor variable. The same model was also refit separately for each of the 13 ADAS sub-scores. For item level analyses, all p-values were FDR corrected for multiple comparisons, with statistical significance as p< 0.05. Analyses were conducted using RStudio (version 2023.03.0-daily+82.pro2), a statistical software package for R programming.

## 3. Results

### 3.1. Population characteristics

Baseline features of the sample are summarized in Table 1. The study population comprised 849 participants, including 656 CN (59% female) and 193 patients with AD (45% female). The mean age of the population was 72 ± 7.51 years. Female CN participants had a lower mean age (70.03 ± 6.51) than male CN participants (72.52 ± 6.63, p<0.001). Similarly, in the AD group, female patients had a lower average age (72.89 ± 8.17) than male patients (75.27 ± 8.36, p=0.05). In both CN and AD groups, females had significantly lower educational levels compared to males (CN: p<0.001, AD: p=0.003). In both CN and AD groups, females had a greater HPF compared to males (p<0.001).

**Table 1.** Demographic and clinical characteristics of study participants.

|  | CN |  |  |  | AD |  |  |  |
| --- | --- | --- | --- | --- | --- | --- | --- | --- |
|  | Total<br>(n=656) | Female<br>(n=389) | Male<br>(n=267) | P value | Total<br>(n=193) | Female<br>(n=80) | Male<br>(n=113) | P value |
| Age, yr, mean<br>(SD) | 71.05<br>(6.67) | 70.03<br>(6.51) | 72.53<br>(6.63) | <0.001 | 74.28<br>(8.34) | 72.89<br>(8.17) | 75.27<br>(8.36) | 0.05 |
| Education, yr,<br>mean (SD) | 16.7<br>(2.36) | 16.4<br>(2.36) | 17.1<br>(2.30) | <0.001 | 15.8<br>(2.61) | 15.1<br>(2.38) | 16.2<br>(2.67) | 0.003 |
| HPF, mean<br>(SD) | 0.86<br>(0.07) | 0.88<br>(0.06) | 0.84<br>(0.07) | <0.001 | 0.72<br>(0.10) | 0.75<br>(0.09) | 0.69<br>(0.10) | <0.001 |
| ADAS-Cog-13,<br>mean<br>(SD) | 8.60<br>(4.37) | 7.54<br>(3.81) | 10.1<br>(4.69) | <0.001 | 31.2<br>(8.19) | 31.7<br>(8.88) | 30.9<br>(7.69) | 0.470 |

### 3.2. ADAS-Cog 13 total score in CN and AD

In the CN group, on average, female participants had a better total ADAS-Cog-13 score (7.54 ± 3.81) compared to male participants (10.1 ± 4.69, p<0.001). Among AD patients, there was no significant differences between the female (31.7 ± 8.88) and male (30.9 ± 7.69, p=0.47) total ADAS-Cog-13 score. In the regression analysis, sex was significantly associated with the total ADAS-Cog-13 score (*β*3= - 2.09 ± 0.33, p<0.0001) in the CN group, indicating that being female was associated with a lower total ADAS-cog-13 score. In the AD group, however, sex was not associated with the task performance (*β*_3_ =2.12 ± 1.19, p=0.076). In the CN group, ADAS-Cog-13 scores were significantly associated with HPF (β_1_ = −0.12 ± 0.03, p<0.0001), age (*β*2 = 0.11 ± 0.03, p<0.001), and educational level (*β*_4_= −0.29 ± 0.07, p<0.0001). In the AD group, regression analysis only demonstrated significant associations between HPF (*β*_1_ = −0.37 ± 0.07, p<0.0001) and age (*β*_2_= −0.16 ± 0.08, p<0.05) with ADAS-Cog-13 scores. The multiple regression model coefficient estimates, standard errors, and p-values for all covariates in regression analyses are reported in Table 2.

**Table 2.**
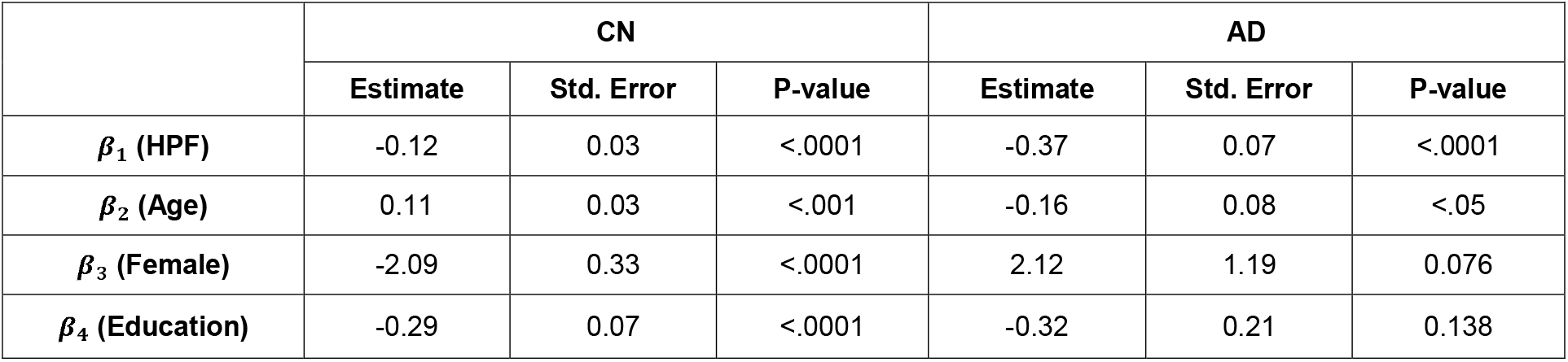
Estimated multilinear regression model coefficients in CN and AD groups.

### 3.3. ADAS-Cog-13 sub-scores in CN and AD

In the CN group, female sex was associated with better performance in word recall (*β*_3_ =3.35 ± 0.08, p<0.001), delayed word recall (*β*_3_ =3.25 ± 0.11, p<0.001), naming (*β*_3_=0.10 ± 0.02, p<0.01), and word recognition (*β*_3_ =2.23 ± 0.12, p<0.001). In AD patients, male sex was associated with better performance in delayed word recall (*β*_3_ =-7.75 ± 0.22, p<0.05). The FDR adjusted p-values for the sub- score specific multiple regression model coefficients are reported in Table 3.

**Table 3.**
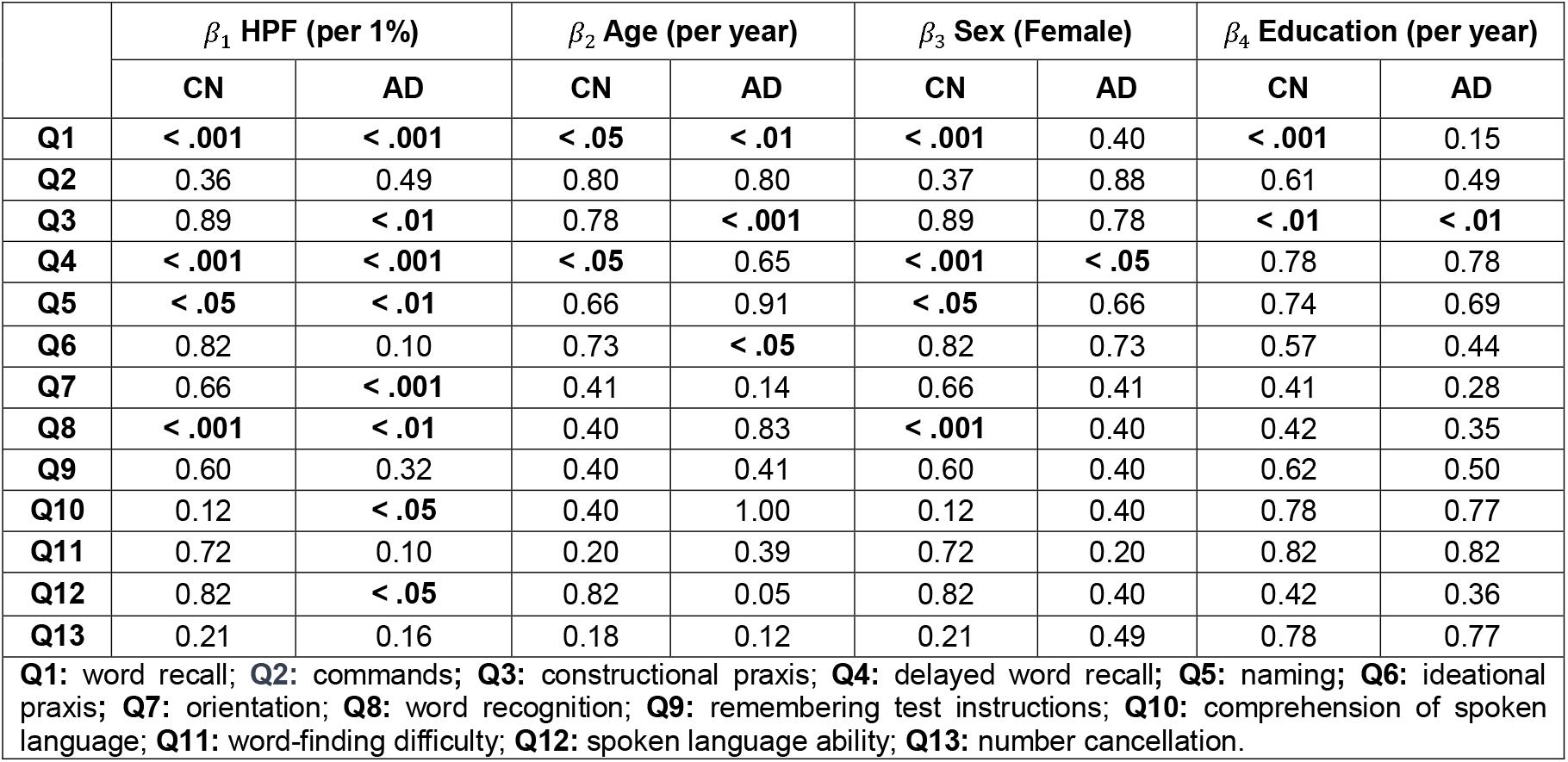
FDR-adjusted p-values for the sub-score specific multilinear regression model coefficients in CN and AD groups.

| | $\beta_1$ HPF (per 1%) | | $\beta_2$ Age (per year) | | $\beta_3$ Sex (Female) | | $\beta_4$ Education (per year) | |
| --- | --- | --- | --- | --- | --- | --- | --- | --- |
|  | CN | AD | CN | AD | CN | AD | CN | AD |
| **Q1** | < .001 | < .001 | < .05 | < .01 | < .001 | 0.40 | < .001 | 0.15 |
| **Q2** | 0.36 | 0.49 | 0.80 | 0.80 | 0.37 | 0.88 | 0.61 | 0.49 |
| **Q3** | 0.89 | < .01 | 0.78 | < .001 | 0.89 | 0.78 | < .01 | < .01 |
| **Q4** | < .001 | < .001 | < .05 | 0.65 | < .001 | < .05 | 0.78 | 0.78 |
| **Q5** | < .05 | < .01 | 0.66 | 0.91 | < .05 | 0.66 | 0.74 | 0.69 |
| **Q6** | 0.82 | 0.10 | 0.73 | < .05 | 0.82 | 0.73 | 0.57 | 0.44 |
| **Q7** | 0.66 | < .001 | 0.41 | 0.14 | 0.66 | 0.41 | 0.41 | 0.28 |
| **Q8** | < .001 | < .01 | 0.40 | 0.83 | < .001 | 0.40 | 0.42 | 0.35 |
| **Q9** | 0.60 | 0.32 | 0.40 | 0.41 | 0.60 | 0.40 | 0.62 | 0.50 |
| **Q10** | 0.12 | < .05 | 0.40 | 1.00 | 0.12 | 0.40 | 0.78 | 0.77 |
| **Q11** | 0.72 | 0.10 | 0.20 | 0.39 | 0.72 | 0.20 | 0.82 | 0.82 |
| **Q12** | 0.82 | < .05 | 0.82 | 0.05 | 0.82 | 0.40 | 0.42 | 0.36 |
| **Q13** | 0.21 | 0.16 | 0.18 | 0.12 | 0.21 | 0.49 | 0.78 | 0.77 |
| **Q1:** word recall; **Q2:** commands; **Q3:** constructional praxis; **Q4:** delayed word recall; **Q5:** naming; **Q6:** ideational praxis; **Q7:** orientation; **Q8:** word recognition; **Q9:** remembering test instructions; **Q10:** comprehension of spoken language; **Q11:** word-finding difficulty; **Q12:** spoken language ability; **Q13:** number cancellation. |  |  |  |  |  |  |  |  |

## 4. Discussion

Our findings indicate that among cognitively normal participants, female sex is associated with significantly better performance on the total ADAS-Cog-13 score compared to males, after adjusting for age, education, and hippocampal volumetric integrity. However, this advantage was not observed in AD patients, where no significant sex differences were found in total scores. For sub-scores, CN females performed significantly better than males in word recall, delayed word recall, naming, and word recognition. In contrast, among AD patients, male sex was associated with better performance in delayed word recall.

Our findings that females in the CN group performed better on the total ADAS-Cog-13 score aligns with prior research showing that females generally outperform males in episodic memory, verbal memory, and processing speed.^26^ Regarding ADAS-Cog assessment, one study on MCI subjects found no significant difference in baseline ADAS-Cog performance between males and females, but female subjects had a greater longitudinal rate of decline in this task.^27^

In other studies, including subjects with MCI and AD, female advantage in verbal memory has been reported, despite similar levels of brain atrophy.^15,28^ This female advantage may mask early symptoms of cognitive decline in women, leading to delayed diagnosis despite comparable levels of AD pathology. In contrast, other studies have reported that in AD patients, men significantly performed better than women in verbal and visuospatial tasks and tests of episodic and semantic memory.^28,29^ This male advantage in AD patients could possibly be due to faster cognitive decline in women once symptomatic.^27^ These differences do not appear to be attributable to any differences in age, education, or dementia severity, but instead, could be related to the reduction of estrogen in postmenopausal women, greater cognitive reserve in men, and the influence of the apolipoprotein E ε4 allele.^29,30^ Similar to these studies, the absence of significant sex differences in the AD group in our study suggests that the progression of AD pathology may eliminate the cognitive advantages observed in females during the cognitively normal stage.

The sub-score analysis further supports this interpretation. Among CN participants, females showed significantly better performance in multiple verbal domains. These results reinforce the notion of a female advantage in verbal memory tasks and highlight the importance of considering sex as a biological variable in cognitive aging research. Interestingly, in the AD group, males outperformed females in delayed word recall, suggesting a possible reversal in sex-related cognitive trajectories as the disease progresses. Again, this reversal may reflect the accelerated decline in memory function observed in females once AD symptoms emerge, a phenomenon reported in earlier studies.^31,32^

These findings have important clinical implications. Standard cognitive assessments, such as the ADAS-Cog-13, may not fully account for sex differences in baseline cognitive abilities, potentially leading to underdiagnosis or delayed diagnosis in females. The superior verbal memory skills in CN females could mask early cognitive decline, reducing the sensitivity of these tools to detect prodromal AD in women. Therefore, incorporating sex-specific norms or adjusting scoring thresholds based on sex may enhance diagnostic accuracy. Future studies are needed to examine how sex influences cognitive decline trajectories and whether cognitive tests can be optimized to improve detection across sexes.

This study has several limitations. All analyses were conducted within a single cohort from the ADNI database, which may limit the generalizability of the findings. Replicating the analysis across multiple cohorts and expanding the sample size would enhance the robustness and reliability of the results. Additionally, to gain deeper insights into sex differences in cognition, it is important to examine other standard cognitive assessment tools.

## Conclusions

These findings underscore the importance of accounting for sex differences in cognitive aging, as they may affect baseline performance and influence the early detection of Alzheimer’s disease.

## Data Availability

All data used in the preparation of this manuscript were obtained from the Alzheimer Disease Neuroimaging Initiative (ADNI) database.

https://adni.loni.usc.edu/

## Acknowledgements

Data collection and sharing for this project was funded by the Alzheimer’s Disease Neuroimaging Initiative (ADNI) (National Institutes of Health Grant U01 AG024904) and DOD ADNI (Department of Defense award number W81XWH-12-2-0012). ADNI is funded by the National Institute on Aging, the National Institute of Biomedical Imaging and Bioengineering, and through generous contributions from the following: AbbVie, Alzheimer’s Association; Alzheimer’s Drug Discovery Foundation; Araclon Biotech; BioClinica, Inc.; Biogen; Bristol-Myers Squibb Company; CereSpir, Inc.; Cogstate; Eisai Inc.; Elan Pharmaceuticals, Inc.; Eli Lilly and Company; EuroImmun; F. Hoffmann-La Roche Ltd and its affiliated company Genentech, Inc.; Fujirebio; GE Healthcare; IXICO Ltd.; Janssen Alzheimer Immunotherapy Research & Development, LLC.; Johnson & Johnson Pharmaceutical Research & Development LLC.; Lumosity; Lundbeck; Merck & Co., Inc.; Meso Scale Diagnostics, LLC.; NeuroRx Research; Neurotrack Technologies; Novartis Pharmaceuticals Corporation; Pfizer Inc.; Piramal Imaging; Servier; Takeda Pharmaceutical Company; and Transition Therapeutics. The Canadian Institutes of Health Research is providing funds to support ADNI clinical sites in Canada. Private sector contributions are facilitated by the Foundation for the National Institutes of Health (www.fnih.org). The grantee organization is the Northern California Institute for Research and Education, and the study is coordinated by the Alzheimer’s Therapeutic Research Institute at the University of Southern California. ADNI data are disseminated by the Laboratory for Neuro Imaging at the University of Southern California.

## References

1. Alzheimer’s disease. Mayo Clinic. https://www.mayoclinic.org/diseases-conditions/alzheimers-disease/symptoms-causes/syc-20350447 (2024, accessed 21 June 2025).

2. Alzheimer’s Association. 2019 Alzheimer’s disease facts and figures. Alzheimer’s & Dementia 2019; 15: 321–387.

3. Lopez-Lee C, Torres ERS, Carling G, et al. Mechanisms of sex differences in Alzheimer’s disease. Neuron 2024; 112: 1208–1221.

4. Castro-Aldrete L, Moser MV, Putignano G, et al. Sex and gender considerations in Alzheimer’s disease: The Women’s Brain Project contribution. Front Aging Neurosci 2023; 15: 1105620.

5. Ferretti MT, Iulita MF, Cavedo E, et al. Sex differences in Alzheimer disease — the gateway to precision medicine. Nature Reviews Neurology. 2018;14(8):457–469. doi:10.1038/s41582-018-0032-9.

6. Zhu D, Montagne A, Zhao Z. Alzheimer’s pathogenic mechanisms and underlying sex difference. Cell Mol Life Sci 2021; 78: 4907–4920.

7. Asperholm M, Nagar S, Dekhtyar S, et al. The magnitude of sex differences in verbal episodic memory increases with social progress: Data from 54 countries across 40 years. PLoS One 2019; 14: e0214945.

8. Tynkkynen J, Chouraki V, van der Lee SJ, et al. Association of branched-chain amino acids and other circulating metabolites with risk of incident dementia and Alzheimer’s disease: A prospective study in eight cohorts. Alzheimers Dement 2018; 14: 723–733.

9. Sundermann EE, Maki PM, Rubin LH, et al. Female advantage in verbal memory: Evidence of sex-specific cognitive reserve. Neurology 2016; 87: 1916–1924.

10. Sundermann EE, Biegon A, Rubin LH, et al. Better verbal memory in women than men in MCI despite similar levels of hippocampal atrophy. Neurology 2016; 86: 1368–1376.

11. Digma LA, Madsen JR, Rissman RA, et al. Women can bear a bigger burden: ante- and post-mortem evidence for reserve in the face of tau. Brain Commun 2020; 2: fcaa025.

12. Vila-Castelar C, Tariot PN, Sink KM, et al. Sex differences in cognitive resilience in preclinical autosomal-dominant Alzheimer’s disease carriers and non-carriers: Baseline findings from the API ADAD Colombia Trial. Alzheimers Dement 2022; 18: 2272–2282.

13. Voyer D, Saint Aubin J, Altman K, et al. Sex differences in verbal working memory: A systematic review and meta-analysis. Psychol Bull 2021; 147: 352–398.

14. Hirnstein M, Stuebs J, Moè A, et al. Sex/Gender Differences in Verbal Fluency and Verbal-Episodic Memory: A Meta-Analysis. Perspect Psychol Sci 2023; 18: 67–90.

15. Sundermann EE, Maki PM, Reddy S, et al. Women’s higher brain metabolic rate compensates for early Alzheimer’s pathology. Alzheimers Dement (Amst) 2020; 12: e12121.

16. Lin KA, Choudhury KR, Rathakrishnan BG, et al. Marked gender differences in progression of mild cognitive impairment over 8 years. Alzheimers Dement (N Y) 2015; 1: 103–110.

17. Holland D, Desikan RS, Dale AM, et al. Higher rates of decline for women and apolipoprotein E epsilon4 carriers. AJNR Am J Neuroradiol 2013; 34: 2287–2293.

18. Kueper JK, Speechley M, Montero-Odasso M. The Alzheimer’s Disease Assessment Scale-Cognitive Subscale (ADAS-Cog): Modifications and Responsiveness in Pre-Dementia Populations. A Narrative Review. J Alzheimers Dis 2018; 63: 423–444.

19. Wang L, Tian T. Gender Differences in Elderly With Subjective Cognitive Decline. Front Aging Neurosci 2018; 10: 166.

20. Goff DC, Zeng B, Ardekani BA, et al. Association of Hippocampal Atrophy With Duration of Untreated Psychosis and Molecular Biomarkers During Initial Antipsychotic Treatment of First-Episode Psychosis. JAMA Psychiatry 2018; 75: 370–378.

21. Tsiknia AA, Sundermann EE, Reas ET, et al. Sex differences in Alzheimer’s disease: plasma MMP-9 and markers of disease severity. Alzheimers Res Ther 2022; 14: 160.

22. NITRC: Automatic registration toolbox: Tool/resource info. NITR Chttps://www.nitrc.org/projects/art/.

23. Skinner J, Carvalho JO, Potter GG, et al. The Alzheimer’s Disease Assessment Scale-Cognitive-Plus (ADAS-Cog-Plus): an expansion of the ADAS-Cog to improve responsiveness in MCI. Brain Imaging Behav 2012; 6: 489–501.

24. Nogueira J, Freitas S, Duro D, et al. Validation study of the Alzheimer’s disease assessment scale-cognitive subscale (ADAS-Cog) for the Portuguese patients with mild cognitive impairment and Alzheimer’s disease. Clin Neuropsychol 2018; 32: 46–59.

25. Siedlecki KL, Falzarano F, Salthouse TA. Examining Gender Differences in Neurocognitive Functioning Across Adulthood. J Int Neuropsychol Soc 2019; 25: 1051–1060.

26. Irvine K, Laws KR, Gale TM, et al. Greater cognitive deterioration in women than men with Alzheimer’s disease: a meta analysis. J Clin Exp Neuropsychol 2012; 34: 989–998.

27. Ryan JJ, Glass Umfleet L, Kreiner DS, et al. Neuropsychological differences between men and women with Alzheimer’s disease. Int J Neurosci 2018; 128: 342–348.

28. Bourzac K. Why women experience Alzheimer’s disease differently from men. Naturecom. Published online April 16, 2025. doi:10.1038/d41586-025-01106-y

29. Kim J, Basak JM, Holtzman DM. The role of apolipoprotein E in Alzheimer’s disease. Neuron 2009; 63: 287–303.

30. Laws KR, Irvine K, Gale TM. Sex differences in cognitive impairment in Alzheimer’s disease. World J Psychiatry 2016; 6: 54–65.

31. Emrani S, Sundermann EE. Sex/gender differences in the clinical trajectory of Alzheimer’s disease: Insights into diagnosis and cognitive reserve. Front Neuroendocrinol 2025; 77: 101184.

32. Levine DA, Gross AL, Briceño EM, et al. Sex Differences in Cognitive Decline Among US Adults. JAMA Netw Open 2021; 4: e210169.

